# Underlying reasons for post-mortem diagnosed lung cancer cases – A robust retrospective comparative study from Hungary (HULC study)

**DOI:** 10.1101/2022.08.08.22278541

**Authors:** Zoltán Kiss, Krisztina Bogos, Lilla Tamási, Gyula Ostoros, Veronika Müller, Nóra Bittner, Veronika Sárosi, Aladár Vastag, Kata Knollmajer, Máté Várnai, Krisztina Kovács, Andrea Berta, István Köveskuti, Eugenia Karamousouli, György Rokszin, Zsolt Abonyi-Tóth, Zsófia Barcza, István Kenessey, András Weber, Péter Nagy, Petra Freyler-Fadgyas, Miklós Szócska, Péter Szegner, Lászlóné Hilbert, Gabriella Branyiczkiné Géczy, György Surján, Judit Moldvay, Zoltán Vokó, Gabriella Gálffy, Zoltán Polányi

**Affiliations:** MSD Pharma Hungary Ltd, Budapest, Hungary; National Korányi Institute of Pulmonology, Budapest, Hungary; Department of Pulmonology, Semmelweis University, Budapest, Hungary; Matrahaza Healthcare Center and University Teaching Hospital, Matrahaza, Hungary; Department of Pulmonology, University of Debrecen, Debrecen, Hungary; Faculty of Medicine, University of Pécs, Pécs, Hungary; Center for Health Technology Assessment, Semmelweis University, Budapest, Hungary; RxTarget Ltd. Szolnok, Hungary; University of Veterinary Medicine Budapest, Hungary; Syntesia Medical Communications Ltd, Budapest, Hungary; 1^st^ Department of Pulmonology, National Korányi Institute of Pulmonology, Semmelweis University, Budapest, Hungary; National Institute of Oncology, National Tumorbiology Laboratory project (NLP-17), Budapest, Hungary; Department of Pathology, Forensic and Insurance Medicine, Semmelweis University, Budapest, Hungary; Cancer Surveillance Branch, International Agency for Research on Cancer, Lyon, France; Department of Anatomy and Histology, University of Veterinary Medicine, Budapest, Hungary; Institute of Oncochemistry, University of Debrecen, Debrecen, Hungary; National Health Insurance Fund, Budapest, Hungary; Institute of Digital Health Sciences, Semmelweis University, Budapest, Hungary; Hungarian Central Statistical Office, Budapest, Hungary; Ministry of Human Resources, Budapest, Hungary; 2nd Department of Pathology, MTA-SE NAP, Brain Metastasis Research Group, Hungarian Academy of Sciences, Semmelweis University, Budapest, Hungary; Pulmonology Hospital Törökbálint, Hungary; Department of Thoracic Surgery, Semmelweis University, Budapest, Hungary

**Keywords:** lung cancer, post-mortem diagnoses, late stage, lung cancer mortality, delayed diagnosis

## Abstract

**Objective:** The Hungarian Undiagnosed Lung Cancer (HULC) study aimed to explore the potential reasons for missed LC diagnosis by comparing healthcare and socio-economic data among patients with post-mortem diagnosed LC with those who were diagnosed with LC during their lives.

**Methods:** This nationwide, retrospective study used the databases of the Hungarian Central Statistical Office (HCSO) and National Health Insurance Fund (NHIF) to identify patients who died between January 1, 2019 and December 31, 2019 and were diagnosed with lung cancer post-mortem (population A) or during their lifetime (population B). Patient characteristics, socio-economic factors, and healthcare resource utilization (HCRU) data were compared between the diagnosed and undiagnosed patient population.

**Results:** During the study period, 8,435 patients were identified from the HCSO database with LC as the cause of death, of whom 1,203 (14.24%) had no LC-related ICD code records in the NHIF database during their lives (post-mortem diagnosed LC population). Post-mortem diagnosed LC patients were significantly older than patients diagnosed while still alive (mean age 71.20 vs. 68.69 years, p<0.001), with a more pronounced age difference among female patients (difference: 4.57 years, p<0.001), and had significantly fewer GP and specialist visits, X-ray and CT scans within 7 to 24 months and 6 months before death, although the differences in GP and specialist visits within 7–24 months did not seem clinically relevant. Patients diagnosed with LC while still alive were more likely to be married (47.62% vs. 33.49%), had higher educational attainment, and had more children, than patients diagnosed with LC post-mortem.

**Conclusions:** Post-mortem diagnosed lung cancer accounts for 14.24% of total lung cancer mortality in Hungary. This study provides valuable insights into patient characteristics, socio-economic factors, and HCRU data potentially associated with a high risk of lung cancer misdiagnosis.

## INTRODUCTION

Every year, 2.1 million people are newly diagnosed with lung cancer (LC) and 1.8 million people die of LC worldwide, making it the most frequently diagnosed cancer and the leading cause of cancer-related mortality [Bray et al., 2018]. Historically, Hungary has always been positioned at the top of LC incidence ranking among European countries [Ferlay 2012, 2018]. However, a number of recent epidemiological studies have consistently reported lower incidence and mortality rates of LC for Hungary, than previous publications [Bogos 2019, Tamási 2021]. These differences may be attributed to differences in data collection methodology and at least partly to the relatively high autopsy rate for hospital deaths in Hungary which leads to a higher likelihood of diagnosing post-mortem LC, than in other countries with lower autopsy rates [European Health Info Gateway, Bogos 2019]. This hypothesis is supported by several studies which suggest that post-mortem diagnosed LC may account for a clinically relevant proportion of all LC cases [McFarlane et al., 1986, Kendrey et al., 1996, Marel et al., 2015, Egerváry et al., 2000]. McFarlane et al. conducted a review of post-mortem reports at a university hospital and found that 28% of primary lung cancers detected during autopsy had not been diagnosed while the patient was still alive [McFarlane et al, 1986]. A Hungarian study based on 2,000 autopsies performed at two university hospitals found that 59% of LC cases were newly detected during autopsy, raising attention to the common misdiagnosis of the disease [Szende B et al., 1996]. In 2000, Egerváry et al. examined 250 consecutive autopsies in a specialized pulmonology center in Hungary and found that the false negative rate for LC diagnosis was 9%, however, Marel et al. reported a much higher proportion of undiagnosed LC (44%) in a 2-year study comparing autopsy findings with clinical data in 2015 [Egerváry et al., 2000, Marel et al., 2015].

Although significant improvements in diagnostic accuracy have likely resulted in a decrease in the rate of LC misdiagnosis since early studies, more recent reports suggest that there is still room for improvement in this field and highlight the need for the identification of factors leading to LC misdiagnosis. In Hungary, the comprehensive database of the National Health Insurance Fund Administration (NHIF) contains data from all patients who are diagnosed with and die of LC, while the database of the Hungarian Central Statistical Office (HCSO) contains cause-specific mortality data for all cancer types including LC based on medical certificates of death. Comparing the two databases allows for the identification of post-mortem diagnosed LC cases and the exploration of factors leading to LC misdiagnosis.

Therefore, the aim of the Hungarian Undiagnosed Lung Cancer (HULC) study was to explore the potential reasons for missed LC diagnoses by comparing healthcare and socio-economic data of patients with post-mortem diagnosed LC and those who were diagnosed with LC during their lives.

## MATERIALS AND METHODS

This retrospective, longitudinal study was based on the NHIF and HCSO databases. The NHIF is a nationwide health insurance system which covers almost 100% of the Hungarian population and collects specific information regarding all in- and outpatient visits, including patients’ ID and ICD-10 codes, as well as prescriptions of reimbursed drugs. Furthermore, the NHIF database records the exact dates of all interventions as well as their recorded ICD-10 codes. The HCSO is a professionally independent, self-managed government office which collects mortality data covering 100% of the Hungarian population. Medical certificates of death are collected from hospitals, general practitioners (GPs), and autopsy departments. Hungarian physicians are the only ones entitled to determining the cause of death, as registered in the HCSO database. HCSO finally defines the underlying cause of death based on WHO guidelines.

The current analysis included patients with a diagnosis of LC (ICD-10 code: C34) who died between January 1, 2019, and December 31, 2019. LC was defined as follows: (i) age above 20 years at the time of diagnosis; (ii) a minimum of two occurrences of the ICD-10 code C34 within more than 30 but less than 365 days between January 1, 2009 and December 31, 2019. To avoid the potential miscoding of LC and ensure that only patients truly involved in LC care would be included, patients with cancer-related ICD-10 codes other than C34 as well as those receiving anticancer therapy other than LC treatment within 6 months before or 12 months after the first occurrence of C34 were excluded if they did not have LC-related histology codes in patient records.

Patients who had died of LC were identified from the HCSO database based on the appearance of the ICD-10 code C34 on death records as the main cause of death or disease that existed at time of death between January 1, 2019, and December 31, 2019. The following two patient populations were established by connecting the HCSO and NHIF databases based on patient IDs: (i) patients fulfilling the inclusion criteria in the NHIF database and having C34 in the HCSO database constituted the **diagnosed and deceased LC population (population B)**; (ii) patients without C34 codes in the NHIF database who had LC as the cause of death in the HCSO database were considered undiagnosed and deceased LC patients (**post-mortem diagnosed LC, population A**). Consequently, the undiagnosed LC population was identified solely from the HCSO database. Despite having no ICD-10 C34 code records in the NHIF database, these patients had NHIF data regarding healthcare recourse use, allowing for the evaluation of healthcare interventions 2 years before the time of death.

The following patient ID-based information was collected from the NHIF: number of physician visits (GP and specialist visits), days of hospitalization, number of chest X-rays and chest CTs. We gained the following information from HCSO databases: marital status, number of children at the time of death, the size of the settlement where the deceased person lived at the time of death, educational status, age, sex, and the type of physician identifying the cause of death.

Patient socioeconomic characteristics and healthcare resource utilization data were compared between the diagnosed and deceased LC population and the undiagnosed and deceased population (post-mortem diagnosed). The percentages of patients having GP or specialist visits or hospitalizations, X-ray, or CT diagnoses within 6 months and 7–24 months before death were also compared between the two groups. Furthermore, we examined the differences in annual GP and specialist visits per capita, the annual number of X-ray or CT diagnoses, and mean days of hospitalization also separately within 6 months and 7–24 months before death. Marital status, number of children at the time of death, educational status, and the size of the settlement and main Hungarian region where the deceased person lived at the time of death were also compared.

The mean age at death, the mean annual number of GP visits, specialist visits, hospital days, X-ray and CT examinations were compared by independent samples t-test. The different aspects of the socio-economic status were compared by Pearson’s chi-squared test. About 920 diagnosed lung cancer patients had no C34 record as the cause of death in the HCSO database, therefore, their socio-economic parameters were not available for our analysis.

All calculations were performed with R version 3.5.2 (20/12/2018). The protocol of our study was approved by the National Ethical Board for Health Research (IV/3047-3 /2021/EKU).

## RESULTS

Between January 1, 2019 and December 31, 2019, altogether 8,435 patients were identified from the HCSO database with LC as the underlying cause of death selected for statistical disclosure, of whom 1,203 (14.24%) had no LC-related ICD code records in the NHIF database (post-mortem diagnosed LC population). In this post-mortem diagnosed LC population, the diagnosis of LC as the main cause of death was established by a pathologist in 842 cases, and LC-related death was reported by other physicians in 361 cases. Only patients with LC as the main cause of death established by a pathologist following autopsy (post-mortem identified LC) were included in further analyses. This patient population was compared to patients who died in 2019 and had LC-related ICD code records in the NHIF database (n=7,545) between 2009 and the time of death (LC diagnosed during life) cases (**Figure 1**).

**Figure 1.**
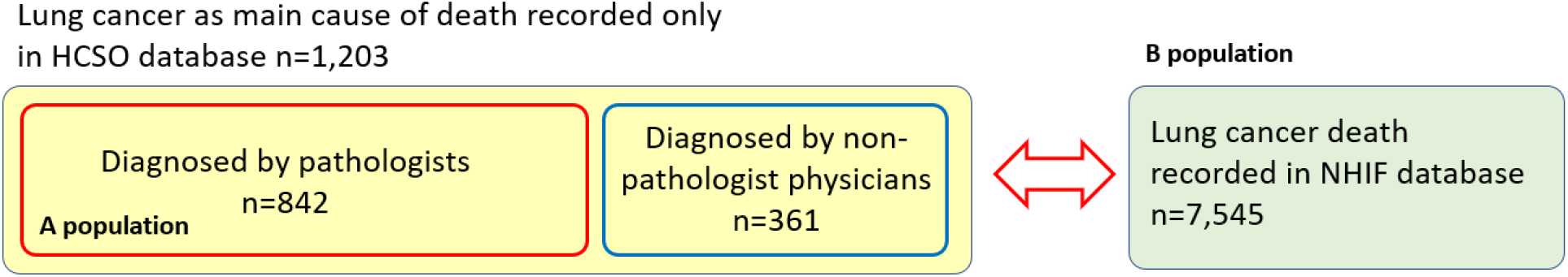
Patient populations included in the HULC study. HULC: Hungarian Undiagnosed Lung Cancer.

The mean age of patients with a post-mortem diagnosis of LC was 71.20 years; 69.59 years among males and 73.77 years among females, all significantly higher than the mean age of patients who were diagnosed with LC during their lives (68.69, 68.33, and 69.20 years, respectively, and p<0.001, p=0.006, and p<0.001, respectively). In the post-mortem diagnosed LC population (population A), 63.27% of females were aged ≥70 years, compared to only 42.66% in population B. There was no relevant difference in the distribution of sex between the two evaluated populations; the proportion of men was 59.81% in the post-mortem diagnosed LC population and 59.36% among those who were diagnosed during their lifetime (**Table 1**).

**Table 1.** Patient characteristics of the study population.

|  | Post-mortem diagnosed deceased patients (A) |  |  | Diagnosed lung cancer patients (B) |  |  |
| --- | --- | --- | --- | --- | --- | --- |
|  | Male | Female | Total | Male | Female | Total |
| No of patients | 518 | 324 | 842 | 4,479 | 3,066 | 7,545 |
| Mean age (SD) | 69.59 (±10.01) | 73.77 (±10.58) | 71.20 (±10.43) | 68.33 (±8.96) | 69.20 (±9.59) | 68.69 (±9.23) |
| Difference A vs B (year, 95%CI; p-value) | 1.26<br>(0.36-2.16; p=0.006) | 4.57<br>(3.37-5.78; p<0.001) | 2.51<br>(1.78-3.25; p<0.001) |  |  |  |
| Distribution of age cohorts (% of total) |  |  |  |  |  |  |
| 20-50 | 15 2.90% | 3 0.93% | 18 2.14% | 124 2.77% | 71 2.32% | 195 2.58% |
| 51-60 | 73 14.09% | 33 10.19% | 106 12.59% | 661 14.76% | 461 15.04% | 1,122 14.87% |
| 61-70 | 211 40.73% | 83 25.62% | 294 34.92% | 1,938 43.27% | 1,226 39.99% | 3,164 41.94% |
| 71≤ | 219 42.28% | 205 63.27% | 424 50.36% | 1,756 39.21% | 1,308 42.66% | 3,064 40.61% |

**Figure 2** shows the mean annual number of GP visits, specialist visits, and hospital days within 7—24 and within 6 months before the time of death in the post-mortem diagnosed (population A) and in the diagnosed and deceased LC population (population B). The majority of post-mortem diagnosed patients had GP visits, specialist visits, or hospitalizations within 24 months before the time of death (92.4%, 92.6%, and 94.4%, respectively).

**Figure 2.**
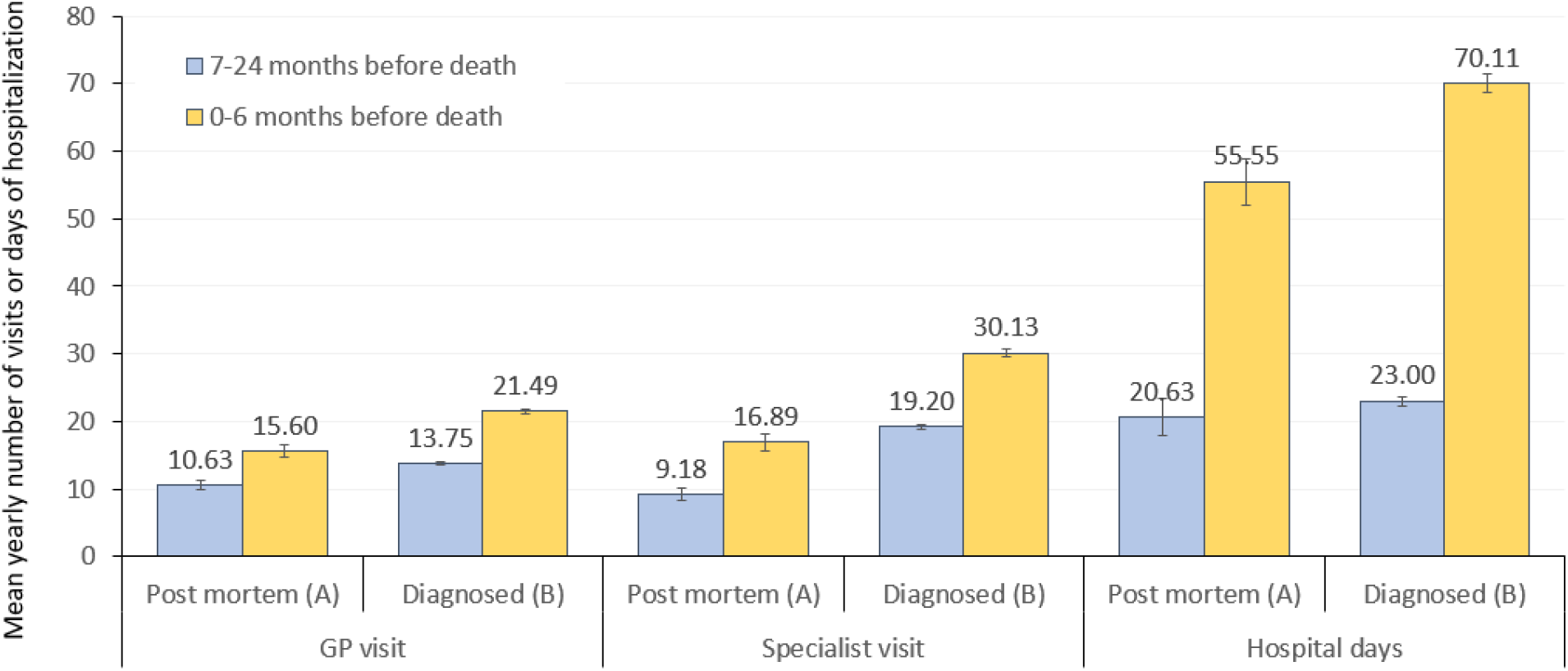
Mean annual number of GP visits, specialist visits, and hospital days within 7–24 months (blue) and 6 months (yellow) before the time of death among patients diagnosed with LC post-mortem (population A) or during their lives (population B). GP: general practitioner; LC: lung cancer.

The mean annual number of GP visits 7–24 months before death was significantly higher among patients who were diagnosed with LC during their lives compared to post-mortem diagnosed LC patients (10.63 vs. 13.75, respectively, p<0.001) (**Supplementary Table 1**). Corresponding numbers within 6 months before death were 15.06 and 21.49 in population A and population B, respectively (p<0.001). The mean annual number of specialist visits was also significantly higher in population B vs. population A both within 7–24 months (19.20 vs. 9.18, p<0.001) and within 6 months before death (30.13 vs. 16.89, p<0.001). The mean annual number of hospital inpatient days was higher but not statistically significantly among diagnosed and deceased LC patients vs. post-mortem diagnosed LC patients within 7–24 months (23.00 vs. 20.63, p=0.094), and significantly higher within 6 months before death (70.11 vs. 55.55 days, p<0.001) (**Figure 2**).

Within 24 months prior to the time of death, 66.0% of post-mortem diagnosed LC patients underwent X-ray examinations and 74.2% had CT scans (any types), compared to 67.0% and 74.6% in patients who were diagnosed during their lives. Within 7 to 24 months before death, the mean annual number of X-ray examinations was 0.44 in the post-mortem diagnosed LC population and 1.37 among those who were diagnosed with LC during their lives; the difference was significant (p<0.001) (**Figure 3, Supplementary Table 1**).

**Figure 3.**
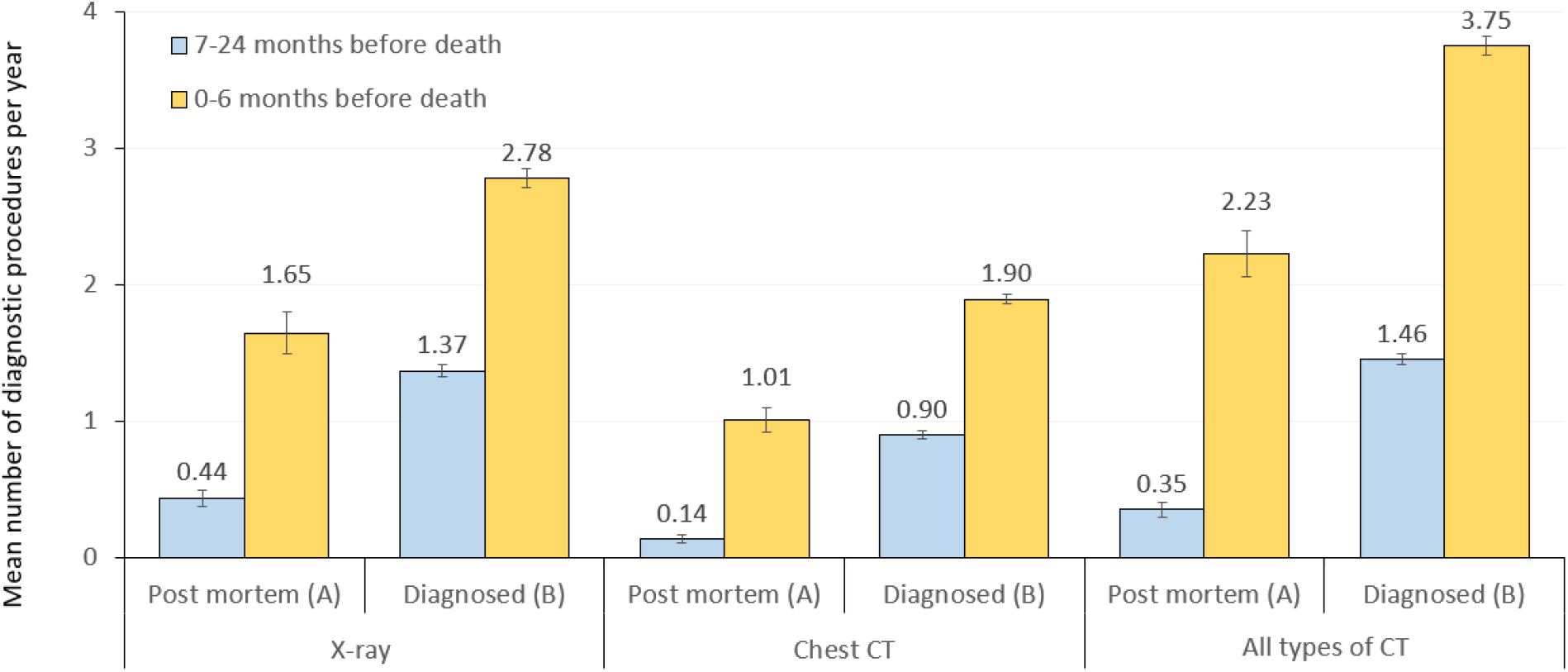
Mean annual number of X-ray, chest CT or all types of CT scans within 7–24 months (blue) and 6 months (yellow) before the time of death among patients diagnosed with LC post-mortem (population A) or during their lives (population B). LC: lung cancer.

Within 6 months before death, the mean annual number of X-ray examinations was 1.65 and 2.78 in population A and population B, respectively (p<0.001). The mean annual number of chest CT scans was also significantly higher in population B vs. population A both within 7–24 months (0.90 vs. 0.14, p<0.001) and within 6 months before death (1.90 vs. 1.01, p<0.001).

As for the socio-economic analysis, the results showed that the percentage of married patients was significantly lower in the post-mortem diagnosed LC population compared to those diagnosed with LC during their lives (33.49% vs. 47.62%, respectively) (**Table 2**). In the post-mortem diagnosed LC population, 23.72% of patients were divorced, 12.28% were single, and 30.51% were widowed at the time of death, compared to 20.40%, 8.81%, and 23.17% among patients diagnosed with LC during their lives (p<0.001 for all parameters). Patients diagnosed with LC post-mortem were significantly more likely to be less educated: 53.27% of population A patients had only primary education (0–8 classes of elementary school), versus 41.94 of patients constituting population B (p<0.001). Post-mortem diagnosed LC patients had fewer children at the time of death: 44.15% had no child (vs. 31.67% in population B, p<0.001), 16.59% had only one child, and 26.46% had 2 children. There was no significant difference between population A and B in terms of settlement size (p=0.485).

**Supplementary Table 1.**
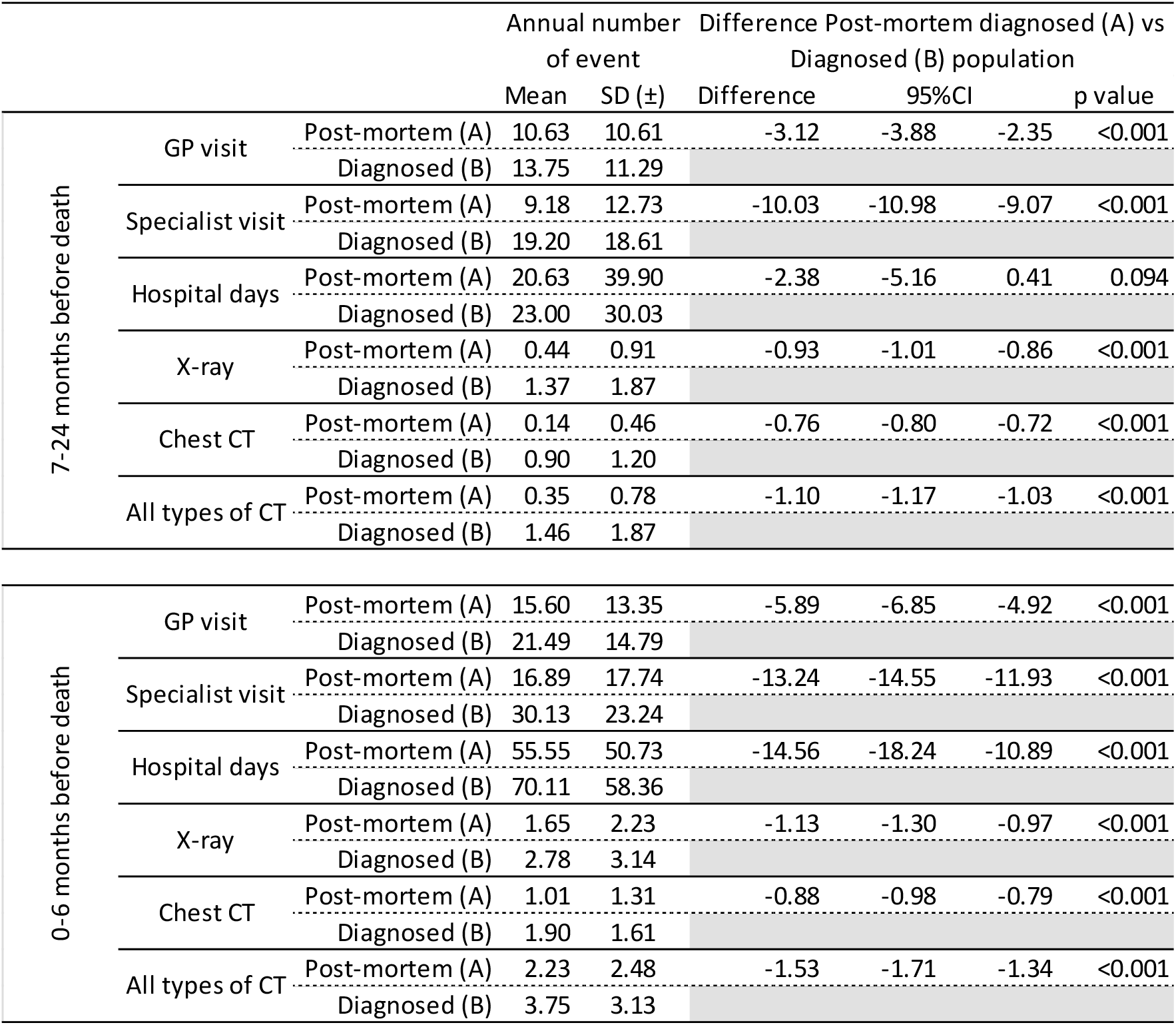
Mean annual number of GP visits, specialist visits, hospitalization days, X-ray examinations, chest CT scans, and all CT scans within 7–24 months and within 6 months before death among patients diagnosed with LC post-mortem (population A) or during their lives (population B). LC: lung cancer; SD: standard deviation.

**Table 2.** Socio-economic status of Hungarian post-mortem diagnosed LC patients and patients who were and diagnosed while still alive. LC: lung cancer.

|  | Number of patients |  | % of population |  | Chi-square |
| --- | --- | --- | --- | --- | --- |
| Main Hungarian regions | Post mortem (A) | Diagnosed (B) | Post mortem (A) | Diagnosed (B) |  |
| Total | 842 | 7,545 |  |  |  |
| **Marrital status** |  |  |  |  |  |
| Divorced | 199 | 1,346 | 23.72% | 20.40% | p<0.001 |
| Single, unmarried | 103 | 581 | 12.28% | 8.81% |  |
| Married | 281 | 3,142 | 33.49% | 47.62% |  |
| Widow | 256 | 1,529 | 30.51% | 23.17% |  |
| **Total identified** | **839** | **6,598** | **99.64%** | **87.45%** |  |
| **Education** |  |  |  |  |  |
| Base education - 0-8 classes | 416 | 2656 | 53.27% | 41.94% | p<0.001 |
| Secondary school | 300 | 3010 | 38.41% | 47.53% |  |
| High school | 65 | 667 | 8.32% | 10.53% |  |
| **Total identified** | **781** | **6,333** | **92.76%** | **83.94%** |  |
| **Number of children** |  |  |  |  |  |
| 0 | 362 | 2043 | 44.15% | 31.67% | p<0.001 |
| 1 | 136 | 1190 | 16.59% | 18.45% |  |
| 2 | 217 | 2205 | 26.46% | 34.19% |  |
| 3+ | 105 | 1,012 | 12.80% | 15.69% |  |
| **Total identified** | **820** | **6,450** | **97.39%** | **85.49%** |  |
| **Size of settlement** |  |  |  |  |  |
| 1 - 9,999 | 354 | 2782 | 42.09% | 42.02% | p=0.485 |
| 10,000 - 49,999 | 184 | 1590 | 21.88% | 24.01% |  |
| 50,000 - | 147 | 1111 | 17.48% | 16.78% |  |
| Budapest | 156 | 1,138 | 18.55% | 17.19% |  |
| **Total identified** | **841** | **6,621** | **99.88%** | **87.75%** |  |

## DISCUSSION

The nationwide, retrospective HULC study provided a unique opportunity to explore the underlying reasons for the significant proportion of post-mortem diagnosed LC patients in the whole LC patient population and to identify potential drivers leading to the delayed diagnosis of such a severe condition.

Significant cross-country differences between autopsy rates limit the international comparison of mortality data. Discrepancies in LC mortality rates reported for Hungary by different studies highlight the importance of conducting analyses which take into account the impact of missed LC diagnoses to allow for the appropriate interpretation of comparative data. As described later in the discussion, the autopsy rate for all deaths has been outstandingly high in Hungary over the past few decades compared to other European countries (35.2% in Hungary vs 12.1% as EU average), as shown by the European Health Information Gateway [European Health Information Gateway]. During autopsy, there is a significant chance for the detection of previously undiagnosed LC. A study in the Czech Republic found a 44% higher incidence of LC in autopsy reports compared to clinical settings [Marel et al., 2015], and other publications also reported a 11% to 28% higher incidence of cancer in autopsy reports than those reported by clinicians [McFarlane et al., 1986, Karwinski et al., 1990]. Previous Hungarian studies also found that a significant proportion of LC was only diagnosed post-mortem [Kendrey et al., 2000, Egerváry et al., 2000]. At the Semmelweis University of Budapest, among patients who died in hospital and underwent autopsy, 59% of lung cancers detected during autopsy were previously undiagnosed and 50% of lung cancers diagnosed during patients’ lives were not confirmed by autopsy [Kendrey et al.]. In a study conducted at the specialized National Korányi Institute of Pulmonology, post-mortem diagnosed lung cancer only accounted for 9% of all lung cancer cases, however, the authors estimated that the false negative rate for the pre-autopsy diagnosis of primary lung cancer was as high as 56% [Egerváry et al.]. A recent study suggests that lung cancer has the highest diagnostic error rates among cancer types (22.5% vs. 2.4% for prostate cancer), which may be attributed to lower-than-recommended screening rates despite public health efforts and resulting delays in diagnosis [Newman-Toker et al., 2020].

The higher the autopsy rate in a country, the higher the chance to detect previously undiagnosed lung cancer. Autopsy rate for all deaths has been outstandingly high in Hungary since 1990, when it was more than double of the mean European Union member (51.0% vs. 22.2%). During the mid-90s and early 2000s, the difference decreased, then in 2019 it was three times higher in Hungary than the European Union average (35.2% vs. 12.1%). While the autopsy rate has been decreasing in most European countries, it remains high in Hungary [European Health Information Gateway] (**Figure 4**).

**Figure 4.**
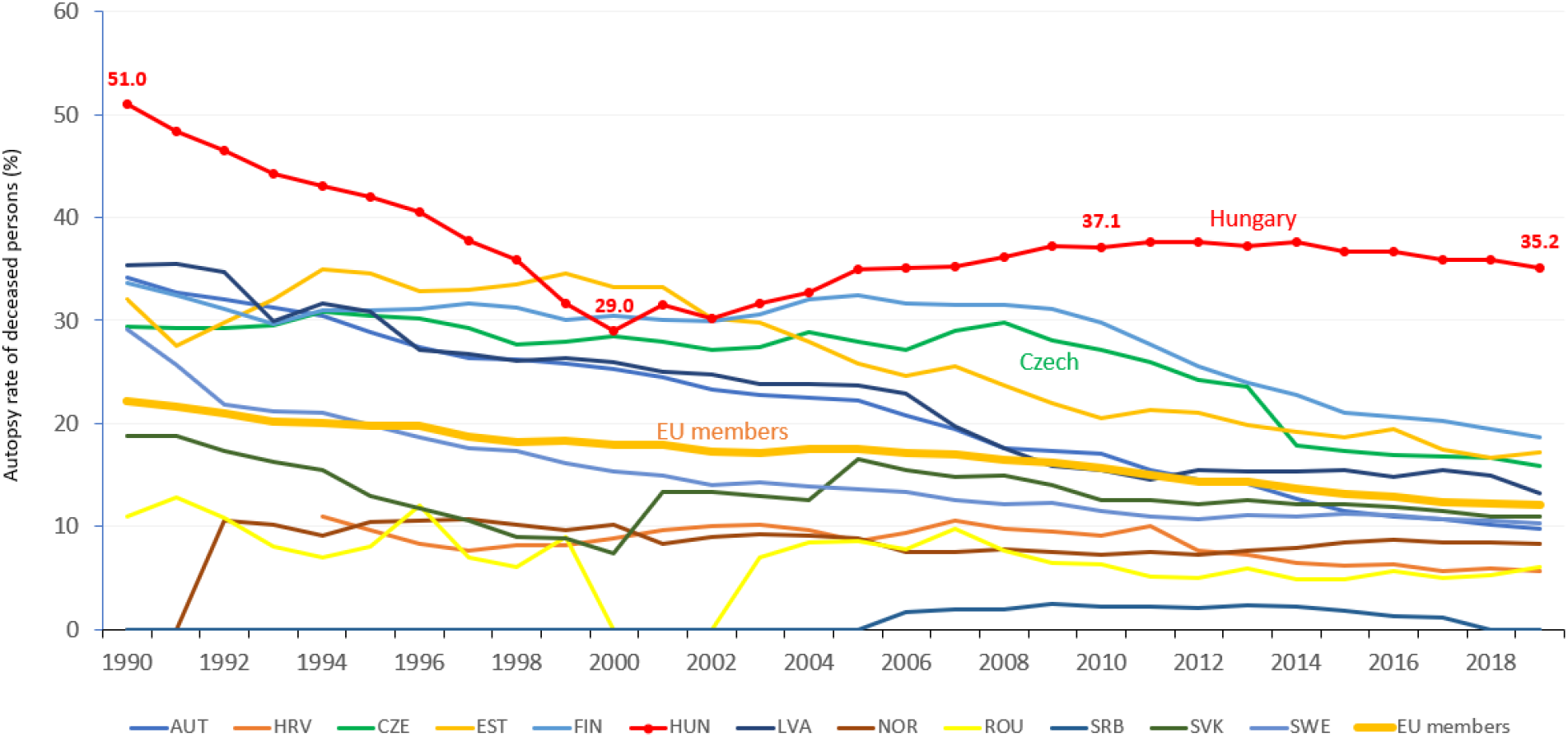
Autopsy rate (%) for all deaths in European countries (where data available). Source: WHO – European Health Information Gateway.

The reason for the high autopsy rate in Hungary is partly a tradition and partly the fact that according to Hungarian law (351/2013. [X. 4.]), autopsy is obligatory for all patients deceased in healthcare facilities unless there are acceptable moral, religious or other grounds for objection. The high rate of autopsy in Hungary, together with a nationwide health insurance database open for research purposes, provides a unique opportunity to investigate the underlying reasons for the missed diagnosis of lung cancer. Connecting the HCSO database, which collects data on the cause of death from hospitals, GPs, and Departments of Pathology, with the NHIF database, which records all healthcare data from almost 100% of the Hungarian population, allowed for the identification of lung cancer patients who were diagnosed post-mortem. We found that post-mortem diagnosed lung cancer accounted for a significant proportion of all lung cancer-related deaths reported for the WHO: 14.24% of patients with lung cancer as the main cause of death had no lung cancer-related ICD code records in the NHIF database, meaning that they had not been diagnosed with lung cancer during their lifetime. Most European countries have considerably lower autopsy rates, than Hungary, which corresponds to a much lower chance of detecting lung cancer post-mortem. Therefore, we examined age-standardized rates of lung cancer mortality in Hungary in 2019 after deducting post-mortem diagnosed lung cancer cases to allow for a better cross-country comparison of mortality rates. Age-standardized mortality was found to be 70.95/100,000 persons in men and 36.54/100,000 persons in women in 2019 using the European Standard Population 1976, which are still among the highest in Europe based on Ferlay’s report from 2018 [Ferlay et al., 2018], and partially contradicts our previous estimates [Bogos et al., 2019, Bogos et al., 2020], which also could be explained by the more restrictive definition of lung cancer. We can conclude that although Hungarian lung cancer mortality rates have been overestimated in cross-country comparisons, but lung cancer still represents a significant burden in Hungary, accounting for one quarter of cancer mortality. These findings underline the importance of developing and implementing more efficient lung cancer management strategies in Hungary in the upcoming years.

Based on our results, more patients die of undiagnosed lung cancer, than the number of patients dying of hepatocellular or bladder cancer in Hungary, which demonstrates the magnitude of the problem. It is therefore crucial to understand the potential factors that may lead to lung cancer misdiagnosis. In our study, post-mortem diagnosed lung cancer patients were significantly older, than patients who were diagnosed with lung cancer during their lives, especially among female patients. Older age and female sex increase the risk of lung cancer misdiagnosis, while the risk of developing cancer including lung cancer also increases with age [White et al., 2014, Tamási et al., 2021]. Limited data are available on age-dependent rates of cancer misdiagnosis, however, several studies have demonstrated age-dependent delays in the lung cancer patient pathway, which may eventually lead to an increase in the risk of cancer misdiagnosis with advancing age [Nadpara et al, 2015].

In our study, a higher level of education was associated with a lower risk of post-mortem lung cancer diagnosis. The awareness of cancer risk, the recognition of cancer-related symptoms, and the willingness to seek help for early diagnosis are higher among people with higher education levels. A recent study conducted in the U.S. showed that the perceived lifetime risk of pancreatic cancer was significantly higher among people with high school education compared to those with lower educational attainment, leading to a higher chance of cancer detection [Underhill et al., 2018]. A recent Hungarian study identified education level as one of the most relevant factors leading to delays in the recognition of testicular cancer-related symptoms and found better survival rates among patients with higher levels of education [Küronya et al., 2021]. In a robust Danish study, the risk of advanced-stage lung cancer was lower in people with higher education levels, but higher in people living alone. In addition, higher levels of education were associated with fewer delays between referral and diagnosis and resulted in a higher proportion of early-stage diagnoses [Dalton et al., 2011]. These findings suggest that low educational attainment may be associated with delayed cancer diagnosis and a higher proportion of post-mortem detected cases, which is confirmed by our current study. Therefore, strategies for the timely mobilization of potentially high-risk lung cancer patients should involve more focus on populations with lower levels of education and individuals living in social isolation.

In our HULC study, marital status and the number of children were also associated with the risk of lung cancer misdiagnosis. Our findings show that married people and those with a higher number of children had a lower risk of lung cancer misdiagnosis, which is in line with previous findings. A recent meta-analysis confirmed the important role of marital status on the early detection of cancer by the higher odds for late-stage breast cancer and malignant melanoma among unmarried people [Buja et al., 2018]. The results highlight the importance of reducing health disparities in the unmarried population. A meta-analysis assessing the impact of socio-economic factors on stage at diagnosis and survival of female patients with gynecological cancers found a 28% increased risk of late-stage diagnosis and a 20% higher risk of death among unmarried female cancer patients, confirming the significant protective effect of marriage against mortality associated with gynecological cancers [Yuan et al., 2021]. The protective effect of marriage may be explained by lifestyle and health-related behaviors, as close partners may recognize cancer-related symptoms in time and encourage their spouse to participate in screening or seek medical advice [Osazuwa-Peters et al., 2019]. Marriage may also reduce risk-taking behavior, increase the chance of optimal diet and exercise, offer protective benefit through family assistance and care, and provide better social support through an extended family network leading to less depression, fatigue, and anxiety [Osborne et al., 2005, Aizer et al., 2013, Buja et al., 2018]. The Danish registry-based study mentioned above also showed an increased risk of advanced-stage lung cancer in people living alone [Dalton et al., 2011]. Marital status and the number of children may therefore be indirectly associated with the risk of post-mortem lung cancer diagnosis as post-mortem detected cancers are usually in advanced stages.

One of the main findings of our HULC study is that patients who were diagnosed with lung cancer only after death had roughly the same annual number of GP and specialist visits within 2 years before the time of death, suggesting that late or missed diagnosis cannot be attributed to the limited access to healthcare resources. Although post-mortem diagnosed lung cancer patients had a slightly lower number of encounters with their GPs (10.63/year vs. 13.75/year), they were having check-ups almost every month. On the other hand, post-mortem diagnosed patients underwent significantly less diagnostic imaging (X-ray and CT scans) within 7–24 months before death compared to patients who were diagnosed with LC during their lives. Considering the similar number of GP visits between the two patient groups, this discrepancy suggests that the suspicion of lung cancer was not raised during GP visits. During the 6 months before death, the number of physician visits, hospitalizations, X-ray examinations, and CT scans showed a marked increase in the post-mortem diagnosed patient group, presumably parallel to the development of cancer-related symptoms. Still, we can assume that the 6-month period prior to death might have been too short to establish the diagnosis of lung cancer, which was only identified as the main cause of death during autopsy. Even if the suspicion of lung cancer was raised, there was a limited time window for confirmatory diagnostic modalities due to the late stage of the disease and probable poor condition of the patient. The results highlight the need for increasing awareness among GPs on the identification of the early signs of the disease. The early detection of lung cancer-related symptoms significantly increases the likelihood of early-stage diagnosis, therefore, GPs have a key role in improving outcomes [Walton et al., 2013]. The CAPER studies (Cancer Prediction in Exeter) demonstrated that cancer-related symptoms were often reported to primary care physicians several months before diagnosis [Hamilton et al., 2009]. Hippisley-Cox et al. developed and validated an algorithm to estimate the risk of having lung cancer and identified independent predictors in primary care settings such as hemoptysis, loss of appetite and weight, cough, decrease in body mass index, increasing deprivation score, smoking, COPD, anemia, and prior cancer [Hippisley-Cox et al., 2011]. These signs and symptoms should raise the suspicion of lung cancer and should be followed by a referral to chest X-ray examination which rarely provides false negative findings for lung cancer [Stapley et al., 2006]. The importance of GP education was demonstrated by a large U.K. study in which the implementation of the 3-year National Awareness and Early Diagnosis Initiative (NAEDI) resulted in an 80.8% increase in the number of community-ordered chest X-rays, an 8.8% increase in the proportion of patients diagnosed at early stages (I-II), and a 9.3% reduction in the proportion of patients diagnosed with late-stage disease (III/IV). The number of X-ray referrals increased after only 3 GP education sessions, which considerably contributed to the overall increase in the diagnosis of early-stage cancer [Kennedy et al., 2018]. The U.K. NAEDI is an illustrative example for a successful nationwide campaign to raise public awareness of the early symptoms of common cancers and could provide guidance for countries with a particularly high burden of lung cancer, such as Hungary.

Patient reluctance to seek medical advice after experiencing cancer-related symptoms increases the risk of late-stage disease at diagnosis [Austoker et al., 2009]. Patient-related delays may derive from the lack of awareness regarding symptoms and risk factors, as well as insufficient awareness about the effectiveness of cancer treatments and the benefits of early detection [MacDonald et al., 2006]. The success of awareness campaigns in terms of early detection and the facilitation of cancer diagnosis has been demonstrated by several studies. In the U.K., a 6-week combined public awareness campaign and GP education programme significantly increased the probability of GP visits due to having a cough and the number of chest X-ray referrals and promoted lung cancer diagnoses among those at the highest risk for lung cancer with high rates of smoking and high levels of social deprivation [Athey et al., 2012]. Another study from the U.K. also found a significant increase in the recognition of persistent cough as a lung-cancer related symptom and a significant increase in the proportion of early-stage lung cancer diagnosis after an awareness campaign [Ironmonger et al., 2015]. In the CHEST Australia Trial, encouraging long-term heavy smokers to seek medical help for respiratory symptoms as early as possible let to a 40% increase in respiratory consultations [Emery et al., 2019]. In a study from Scotland, a one-time consultation with a nurse at the GP’s office and a self-help manual resulted in much earlier GP visits after the development of chest symptoms [Smith et al., 2013]. In summary, the implementation of awareness campaigns and GP educational programmes has the potential to decrease the proportion of late-stage and post-mortem lung cancer diagnoses in Hungary, especially in high-risk patient populations.

Our retrospective study has certain strengths and limitations. We were able to explore the underlying reasons for post-mortem lung cancer diagnoses in a robust patient population due to the high autopsy rate of Hungary and strict reporting requirements towards the HCSO as well as the comprehensive nature of the NHIF database covering almost 100% of the Hungarian population. In addition, the two databases allowed for the analysis of data within 2 years prior to the time of death both in the diagnosed and undiagnosed lung cancer population. This included the analysis of socio-economic factors which were found to have an important role in late diagnosis. On the other hand, we were not able to provide detailed information on the stage of lung cancer at the time of diagnosis or ECOG performance status, and we did not have any data regarding the histology of lung cancer in the post-mortem diagnosed patient population.

## CONCLUSIONS

In conclusion, patients with a post-mortem diagnosis of lung cancer were older, had lower educational attainment, and were more likely to be single compared to patients who were diagnosed with lung cancer during their lives. Elderly unmarried women with a low level of education and fewer children at the time of death were at the highest risk for the misdiagnosis of lung cancer during their lives. However, post-mortem diagnosed lung cancer patients also had access to the healthcare system as demonstrated by the number of GP and specialist visits within 2 years before death, although these encounters did not lead to the detection of lung cancer-related signs and symptoms as shown by the low number of chest X-rays and CT scans performed before death. The findings highlight the importance of public and GP awareness campaigns which may facilitate earlier diagnosis and decrease the proportion of late-stage and post-mortem diagnosed lung cancer, especially among at-risk populations.

## Data Availability

Data sharing is available for the supplementary tables and figures, furthermore the authors are happy to share all available data from the study.

## ACKNOWLEDGEMENTS

We would like to thank the colleagues of NHIF and Hungarian Central Statistical Office to support the connection of their database to identify post-mortem diagnosed lung cancer for 2019. We would also like to thank the NHIF for providing a comprehensive dataset for our analysis, and Zsófia Barcza of Syntesia Medical Communications for medical writing support.

## Ethics approval

Approval was obtained from the Central Ethical Committee of Hungary (IV/3047- 3 /2021/EKU).

## Consent to participate

The study is based on anonymized data collected for financial purposes by the NHIF of Hungary, thus it does not include images or any other personal data that may be used to identify any person. This document has been prepared in part using the Hungarian Central Statistical Office’s lung cancer mortality dataset and in part using the National Health Insurance Fund’s provider reports. The calculations contained in this document and the conclusions drawn from them are solely the intellectual property of the authors.

## Consent to publish

Not applicable.

## Code availability

Not applicable.

## REFERENCES

1. Bray, F, Ferlay, J, Soerjomataram, I, Siegel, RL, Torre, LA, Jemal, A, et al. Global Cancer Statistics 2018: GLOBOCAN Estimates of Incidence and Mortality Worldwide for 36 Cancers in 185 Countries. CA: A Cancer J Clinicians (2018) 7068(46):313394–424. 2018 Jul. doi:10.3322/caac.21492

2. Ferlay, J, Steliarova-Foucher, E, Lortet-Tieulent, J, Rosso, S, Coebergh, JWW, Comber, H, et al. Cancer Incidence and Mortality Patterns in Europe: Estimates for 40 Countries in 2012. Eur J Cancer (2013) 49:1374–403. doi:10.1016/j.ejca.2012.12.027

3. Ferlay, J., Colombet, M., Soerjomataram, I., Dyba, T., Randi, G., Bettio, M., Gavin, A., Visser, O., & Bray, F. (2018). Cancer incidence and mortality patterns in Europe: Estimates for 40 countries and 25 major cancers in 2018.

4. Bogos, K., Kiss, Z., Gálffy, G., Tamási, L., Ostoros, G., Müller, V., Urbán, L., Bittner, N., Sárosi, V., Vastag, A., Polányi, Z., Nagy-Erdei, Z., Vokó, Z., Nagy, B., Horváth, K., Rokszin, G., Abonyi-Tóth, Z., & Moldvay, J. (2019). Revising Incidence and Mortality of Lung Cancer in Central Europe: An Epidemiology Review From Hungary. Frontiers in oncology, 9, 1051. https://doi.org/10.3389/fonc.2019.01051

5. Tamási, L., Horváth, K., Kiss, Z., Bogos, K., Ostoros, G., Müller, V., Urbán, L., Bittner, N., Sárosi, V., Vastag, A., Polányi, Z., Nagy-Erdei, Z., Daniel, A., Nagy, B., Rokszin, G., Abonyi-Tóth, Z., Moldvay, J., Vokó, Z., & Gálffy, G. (2021). Age and Gender Specific Lung Cancer Incidence and Mortality in Hungary: Trends from 2011 Through 2016. Pathology oncology research : POR, 27, 598862. https://doi.org/10.3389/pore.2021.598862

6. World Health Organization – European Health Information Gateway. Autopsy rate (%) for all deaths. Available at: https://gateway.euro.who.int/en/indicators/hfa_545-6410-autopsy-rate-for-all-deaths/ [accessed April 15, 2022]

7. McFarlane MJ, Feinstein AR, Wells CK. Clinical features of lung cancers discovered as a postmortem “surprise”. Chest. 1986 Oct;90(4):520–3.

8. Marel M, Koubkova L, Kovarikova Z, Grandcourtova A, Petrik F, Hroudova H, Capkova L, Kodet R, Fila L. Lung cancer, pulmonary emphysema and pleural effusion: An autopsy study. Biomed Pap Med Fac Univ Palacky Olomouc Czech Repub. 2015 Dec;159(4):642–7.

9. Szende, B., Kendrey, G., Lapis, K., Roe, F. J., & Lee, P. N. (1996). Accuracy of admission and clinical diagnosis of tumours as revealed by 2000 autopsies. European journal of cancer (Oxford, England : 1990), 32A(7), 1102–1108.

10. Egerváry M, Szende B, Roe FJ, Lee PN. Accuracy of clinical diagnosis of lung cancer in Budapest in an institute specializing in chest diseases. Pathol Res Pract. 2000;196(11):761–6.

11. Karwinski B, Svendsen E, Hartveit F. Clinically undiagnosed malignant tumours found at autopsy. APMIS. 1990 Jun;98(6):496–500.

12. Newman-Toker DE, Wang Z, Zhu Y, Nassery N, Saber Tehrani AS, Schaffer AC, Yu-Moe CW, Clemens GD, Fanai M, Siegal D. Rate of diagnostic errors and serious misdiagnosis-related harms for major vascular events, infections, and cancers: toward a national incidence estimate using the “Big Three”. Diagnosis (Berl). 2020 May 14;8(1):67–84.

13. Bogos K, Kiss Z, Gálffy G, Tamási L, Ostoros G, Müller V, Urbán L, Bittner N, Sárosi V, Vastag A, Polányi Z, Nagy-Erdei Z, Vokó Z, Nagy B, Horváth K, Rokszin G, Abonyi-Tóth Z, Barcza Z, Moldvay J. Lung Cancer in Hungary. J Thorac Oncol. 2020 May;15(5):692–699.

14. White MC, Holman DM, Boehm JE, Peipins LA, Grossman M, Henley SJ. Age and cancer risk: a potentially modifiable relationship. Am J Prev Med. 2014 Mar;46(3 Suppl 1):S7–15.

15. Nadpara P, Madhavan SS, Tworek C. Guideline-concordant timely lung cancer care and prognosis among elderly patients in the United States: A population-based study. Cancer Epidemiol. 2015 Dec;39(6):1136–44.

16. Underhill M, Hong F, Lawrence J, Blonquist T, Syngal S. Relationship between individual and family characteristics and psychosocial factors in persons with familial pancreatic cancer. Psychooncology. 2018 Jul;27(7):1711–1718.

17. Küronya Z, Fröhlich G, Ladányi A, Martin T, Géczi L, Gyergyai F, Horváth O, Kiszner G, Kovács Á, Dienes T, Lénárt E, Nagyiványi K, Szarvas T, Szőnyi M, Tóth A, Biró K. Low socioeconomic position is a risk factor for delay to treatment and mortality of testicular cancer patients in Hungary, a prospective study. BMC Public Health. 2021 Sep 19;21(1):1707. doi: 10.1186/s12889-021-11720-w.

18. Dalton SO, Frederiksen BL, Jacobsen E, Steding-Jessen M, Østerlind K, Schüz J, Osler M, Johansen C. Socioeconomic position, stage of lung cancer and time between referral and diagnosis in Denmark, 2001-2008. Br J Cancer. 2011 Sep 27;105(7):1042–8.

19. Buja A, Lago L, Lago S, Vinelli A, Zanardo C, Baldo V. Marital status and stage of cancer at diagnosis: A systematic review. Eur J Cancer Care (Engl). 2018 Jan;27(1). doi: 10.1111/ecc.12755.

20. Yuan R, Zhang C, Li Q, Ji M, He N. The impact of marital status on stage at diagnosis and survival of female patients with breast and gynecologic cancers: A meta-analysis. Gynecol Oncol. 2021 Sep;162(3):778–787.

21. Osazuwa-Peters N, Christopher KM, Cass LM, Massa ST, Hussaini AS, Behera A, Walker RJ, Varvares MA. What’s Love Got to do with it? Marital status and survival of head and neck cancer. Eur J Cancer Care (Engl). 2019 Jul;28(4):e13022. doi: 10.1111/ecc.13022.

22. Osborne C, Ostir GV, D. X, Peek MK, Goodwin JS. The influence of marital status on the stage at diagnosis, treatment, and survival of older women with breast cancer. Breast Cancer Res Treat. 2005 Sep;93(1):41–7.

23. Aizer AA, Chen MH, McCarthy EP, Mendu ML, Koo S, Wilhite TJ, Graham PL, Choueiri TK, Hoffman KE, Martin NE, Hu JC, Nguyen PL. Marital status and survival in patients with cancer. J Clin Oncol. 2013 Nov 1;31(31):3869–76.

24. Walton L, McNeill R, Stevens W, Murray M, Lewis C, Aitken D, Garrett J. Patient perceptions of barriers to the early diagnosis of lung cancer and advice for health service improvement. Fam Pract. 2013 Aug;30(4):436–44.

25. Hamilton W. The CAPER studies: five case-control studies aimed at identifying and quantifying the risk of cancer in symptomatic primary care patients. Br J Cancer. 2009 Dec 3;101 Suppl 2(Suppl 2):S80–6.

26. Hippisley-Cox J, Coupland C. Identifying patients with suspected lung cancer in primary care: derivation and validation of an algorithm. Br J Gen Pract. 2011 Nov;61(592):e715–23.

27. Stapley S, Sharp D, Hamilton W. Negative chest X-rays in primary care patients with lung cancer. Br J Gen Pract. 2006 Aug;56(529):570–3.

28. Kennedy MPT, Cheyne L, Darby M, Plant P, Milton R, Robson JM, Gill A, Malhotra P, Ashford-Turner V, Rodger K, Paramasivam E, Johnstone A, Bhartia B, Karthik S, Foster C, Lovatt V, Hewitt F, Cresswell L, Coupland VH, Lüchtenborg M, Jack RH, Moller H, Callister MEJ. Lung cancer stage-shift following a symptom awareness campaign. Thorax. 2018 Dec;73(12):1128–1136.

29. Austoker J, Bankhead C, Forbes LJ, Atkins L, Martin F, Robb K, Wardle J, Ramirez AJ. Interventions to promote cancer awareness and early presentation: systematic review. Br J Cancer. 2009 Dec 3;101 Suppl 2(Suppl 2):S31–9. doi: 10.1038/sj.bjc.6605388.

30. Macdonald S, Macleod U, Campbell NC, Weller D, Mitchell E. Systematic review of factors influencing patient and practitioner delay in diagnosis of upper gastrointestinal cancer. Br J Cancer. 2006 May 8;94(9):1272–80.

31. Athey VL, Suckling RJ, Tod AM, Walters SJ, Rogers TK. Early diagnosis of lung cancer: evaluation of a community-based social marketing intervention. Thorax. 2012 May;67(5):412–7.

32. Ironmonger L, Ohuma E, Ormiston-Smith N, Gildea C, Thomson CS, Peake MD. An evaluation of the impact of large-scale interventions to raise public awareness of a lung cancer symptom. Br J Cancer. 2015 Jan 6;112(1):207–16.

33. Emery JD, Murray SR, Walter FM, Martin A, Goodall S, Mazza D, Habgood E, Kutzer Y, Barnes DJ, Murchie P. The Chest Australia Trial: a randomised controlled trial of an intervention to increase consultation rates in smokers at risk of lung cancer. Thorax. 2019 Apr;74(4):362–370.

34. Smith S, Fielding S, Murchie P, Johnston M, Wyke S, Powell R, Devereux G, Nicolson M, Macleod U, Wilson P, Ritchie L, Lee AJ, Campbell NC. Reducing the time before consulting with symptoms of lung cancer: a randomised controlled trial in primary care. Br J Gen Pract. 2013 Jan;63(606):e47–54.

